# Complex Chronic Diseases Program: Program Description & Health Outcomes Assessment from a Clinical Data Registry

**DOI:** 10.1101/2024.05.25.24307912

**Authors:** Emily Meagher, Tianna Magel, Travis Boulter, Carola Muñoz, Nicole Prestley, Wee-Shian Chan, Cassandra Bryden, Luis Nacul

## Abstract

**Introduction:** The Complex Chronic Diseases Program (CCDP) was funded by the BC Ministry of Health to address gaps in health services provision for Complex Chronic Diseases (CCDs). The Program offers medical interventions and education on self-management for people with Myalgic Encephalomyelitis/Chronic Fatigue Syndrome (ME/CFS), Fibromyalgia (FM), and Chronic Lyme-Like Syndrome (CLLS). The CCDP Data Registry was created in 2017 for monitoring participant outcomes and program evaluation.

**Methods:** This research outlined the CCDP model of care and analyzed patient-reported questionnaires and clinical data collected longitudinally from consented CCDP Data Registry participants spanning June 2017 through September 2022. T-tests and linear regression modelling were conducted to ascertain changes in symptom presentation across program involvement. These analyses specifically targeted data at the following time points: baseline, 6 months follow-up, and discharge.

**Results:** Data reported in this study represented 668 eligible participants from the 1-year Program. Demographically, the average age was 49 years old (SD=13), 90% were women (n=557), 54% were diagnosed with ME/CFS and FM (n=360), and 36% reported being on long term illness/disability (n=219). Between baseline and discharge, participants with ME/CFS and FM reported improvements in overall physical and mental health, but no significant improvement in other symptom domains such as sleep, fatigue, and pain. The duration of disease at baseline was only related to sleep quality. The previous, more individualized model of care showed better mental health outcomes at 6-months follow-up.

**Discussion:** This analysis showed that CCDP patients experienced relatively severe and persistent symptom presentations. Participants involved in the Program experienced some health benefits at discharge, but further research and interventions are needed to optimize health outcomes. The reliance on self-report of symptoms and the absence of a control group without intervention limit the significance of these findings. A Strategic Direction Plan was developed by the CCDP which emphasized improved training, decentralized services, fast-tracking of eligible individuals, and enhanced education for better patient outcomes.

## Introduction

Complex Chronic Diseases (CCDs) affect approximately 2.9% of Canadians, or around 855,000 people aged 12 years or older [1]. CCDs encompass Fibromyalgia (FM), Myalgic Encephalomyelitis/Chronic Fatigue Syndrome (ME/CFS), and Chronic Lyme-Like Syndrome (CLLS). ME/CFS manifests as persistent, disabling fatigue and general malaise (feeling unwell, flu-like symptoms etc.), worsened by physical activity, and involving sleep dysfunction and impaired cognitive function typically presenting as ‘brain fog’ or difficulty concentrating, and processing information [2]. FM is characterized by widespread chronic pain persisting for at least three months, with patients frequently experiencing increased sensitivity to pain (hyperalgesia) or pain from typically non-painful stimuli (allodynia)[3]. CLLS, though the least understood among these, shares many symptoms with ME/CFS and other post-viral conditions, as well as with FM [4,5]. Patients with CCDs have persistent and debilitating symptoms [6] coupled with functional impairment [7–9]. They often have great difficulty accessing consistent medical care in a coordinated fashion within the healthcare system, frequently leaving them to advocate for themselves [10].

### Complex Chronic Diseases Program – Background and History

The Complex Chronic Diseases Program (CCDP) was the first tertiary care service of its kind in Canada and now one of only three centres dedicated to the care of people with CCDs. The CCDP began operating with funding from the British Columbia (BC) Ministry of Health at BC Women’s Hospital + Health Centre in Vancouver, Canada in 2012 to bridge gaps in health services provision. The Program provides comprehensive evidence-informed care to adults living with CCDs and is delivered by an interprofessional team of healthcare providers. The team’s composition includes representation from internal, family and occupational medicine, infectious diseases, nursing, social work, physiotherapy, occupational therapy, counselling, naturopathy, and other allied health disciplines. The CCDP model of care includes assessments to make or confirm diagnoses, development of treatment plans, education for patients and care givers, as well as linkages with community and primary care resources.

At inception, there were limited models to inform the BC-based CCDP model of care; as such, the Program adopted an ongoing quality improvement model to continuously respond to the needs of the CCDP’s patient population. The CCDP and BC Women’s Hospital leadership, clinicians, and patient partners worked in collaboration to apply LEAN methodology, a type of change management framework, to optimize service quality and clinic flow [11]. These changes were successful in increasing the number of new patients seen.

The CCDP engages in knowledge mobilization activities by providing education and training to physicians, other health care providers, as well as student learners locally and provincially, on complex chronic disease diagnosis and management. The CCDP also supports continuous quality improvement initiatives and conducts clinical and epidemiological research aimed at enhancing the understanding of CCDs, investigating novel tests and interventions, and improving the quality of care and service delivery to patients. An iterative approach has been taken to the CCDP’s model, in which care methods are continually revised and improved based on current best evidence, clinician’s experience and patient partner input. Adults with symptoms, or a diagnosis of any of the complex chronic diseases mentioned above, are eligible for referral to the Program.

From 2012 to 2019, the CCDP focused on a one-to-one consultative model of care, using individual appointments with clinicians while also providing educational materials and a select set of education groups. However, by late 2018, the Program’s rapidly growing waitlist prompted a necessary review of the model of care to better respond to demand. In January of 2019, the CCDP moved to a mixed model of care, with the continuation of individual (physician-led) longitudinal consultations and expansion of the group-based program, the latter led by the interprofessional team. Key changes included expanding educational resources, group discussions and education sessions, and increased virtual care delivery. The number of one-to-one visits with nursing and allied health staff was reduced and targeted to patients with needs or challenges that make group participation unfeasible, such as severe disease symptoms, cognitive or communication issues, or language barriers. Despite these adjustments, all patients continued to receive evaluations and regular individual consultations with CCDP medical specialists, along with referrals to additional clinical services when necessary.

In 2022, the CCDP prepared a Strategic Direction Plan outlining the need for further program development [12]. Improving timely access to care, depends not only on significant increases in knowledgeable providers, but also on the re-structuring of care to be provided more efficiently and according to population and individual needs. The approach outlined in the Strategic Direction Plan focused on: (i) re-imagining the service delivery model, with expansion of care provision at primary care, and the potential development of one or two satellite clinics in different regions of the Province, (ii) community and patient partnerships, and (iii) education of health professionals and the public, training, and knowledge translation. These changes require developing capacity across the continuum of health care across the Province, from primary to specialized care.

### Complex Chronic Diseases Program – Patient Flow

The CCDP’s model of care and patient flow as of August 2024 is summarized in Figure 1, showing two parallel streams of patient care activities. The intake medical assessment includes confirmation of diagnosis and eligibility for inclusion in the Program. Once in the Program, the patient may follow either, or both, the “Medical stream” (with emphasis on medical treatment and review) or “Group stream” (with emphasis on group activities focused on education and patient empowerment for self-management). Group stream patients are given four case coordination appointments, each approximately three months apart. Patients are offered introductory videos covering information about the CCDP and are provided with educational content on their disease and key aspects of its management. The purpose of these appointments is to support patients with program navigation, questions they may have, choosing elective groups (based on their health priorities), and to support them with medical appointments and discharge.

**Figure 1.**
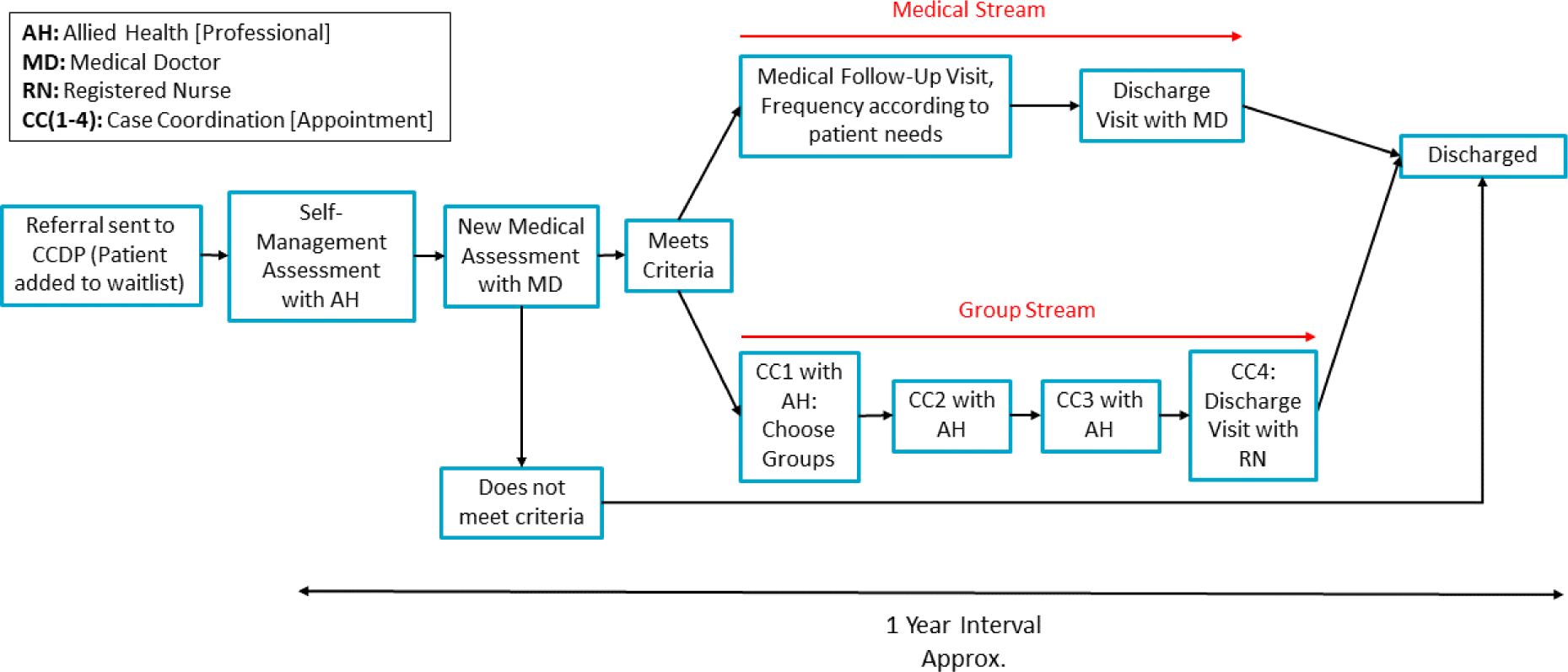
Typical progression for patients involved in the CCDP from referral to discharge.

Upon completion of the Program, patients are discharged and offered the option of joining monthly CCDP alumni network events. The range of alumni topics are intended to support patients with their journey of finding and continuing strategies for self-management that work for them. The CCDP also co-hosts Disability Alliance of BC events which covers different disability resources such as the Canadian Pension Plan Disability Benefits, disability tax credits, and disability assistance. These events are open to the broader community, not just CCDP alumni.

### Patient Education and Self-Management

Patient education is key to the CCDP model of care. A variety of resources and services are offered to patients through group activities. Education is centered on self-management techniques, navigating the healthcare system, workforce and school involvement, social services, family relationships, and symptom management. Each patient is offered a large variety of groups intended to familiarize them with important aspects of their symptom management. These elective groups loosely encompass the major categories of symptom management for CCDs. Groups differ in duration and format, with some groups involving participation in a video-conference meeting while others require the patient to review information packages or videos in addition to attending a virtual visit.

### Clinical Management

Diagnosis is based on clinical criteria, including either the 2003 Canadian Consensus Criteria [2] or the 2015 Institute of Medicine Diagnostic Criteria [13] for diagnosis confirmation of ME/CFS and the 2016 Diagnostic Criteria for Fibromyalgia [14,15]. For CLLS, the diagnosis requires reliable laboratory evidence of Lyme infection in the past and symptoms akin to ME/CFS. The CCDP uses its own guidelines for clinical management, which are available on its website [16]. The clinical team, as appropriate, also follows the principles of the National Institute for Health and Care Excellence (NICE) [17] and the European Network on Myalgic Encephalomyelitis / Chronic Fatigue Syndrome (EUROMENE) guidance on ME/CFS [18]. In accordance with this guidance, the CCDP uses pacing and energy management as a key strategy for symptom control and recovery.

### Partnership work

The CCDP interdisciplinary and leadership team works collaboratively with diverse stakeholders, including patient partners, health care decision makers, and research partners. These relationships enable the CCDP to evolve with the needs of patients, and design new systems. Collaborative endeavors like the CCDP Community Advisory Committee allow real-world perspectives from patients and caregivers to be integrated into programming and operations. Partnership and patient-engagement are also integrated in CCDP research whenever possible, including its activities as part of the Interdisciplinary Canadian Collaborative Myalgic Encephalomyelitis (ICanCME) Research Network and patient engagement research, such as a needs assessment for people with ME/CFS in the province of British Columbia [19].

### Complex Chronic Diseases Program – Data Registry

To support data-driven program evaluation and foster continuous quality improvement and clinical research, the CCDP Data Registry was launched in 2017. This registry functions as an instrument for characterizing the clinic population, monitoring patient outcomes, facilitating future research, and providing program evaluation. The need for the registry arose from the limitations of the existing electronic medical records systems at the hospital, which did not allow for the systematic longitudinal collection of patient information from those who were referred to the CCDP. Gathering and summarizing critical information such as diagnoses, disease course, severity and duration of illness, patient histories, and clinical outcomes data was previously resource intensive and cumbersome. The CCDP Data Registry facilitates the assessment of symptom progression through various questionnaires administered as patients navigate the Program, continuing until three to six months post-program completion. The creation of the CCDP Data Registry presents a unique opportunity to characterize BC patients with CCDs that are seen by this provincial referral service.

In collaboration with clinical staff, patients, and researchers, a suite of questionnaires was identified to serve dual clinical and research purposes. Patient partners reviewed the proposed questionnaires to assess feasibility of completion. Additional data sources were ascertained from clinical chart information. One of the main objectives of the Data Registry was to characterize the CCDP patient population. To achieve this, a comprehensive demographics questionnaire was created to capture details not collected during routine clinical care.

In October 2022, due to funding restrictions, the enrollment of new CCDP Data Registry participants was paused and only participants previously registered continued taking part in follow-up data collection (6-Months, Discharge, and 6-Months Post-Discharge).

## Methods

### Registry Participants

The inclusion criteria for the Data Registry were: (a) newly referred and contacted patients of the CCDP; (b) able to read and understand English; (c) 18 years of age or older; (d) informed consent to be part of the Registry. Exclusion criteria for the study were: (a) not completing the Standardized Questionnaires prior to their intake assessment; (b) the patient being found not eligible for participation in the CCDP clinical program, e.g. for not meeting diagnosis of one of the CCDs managed at CCDP.

After approval by the research ethics board at the University of British Columbia (Vancouver, BC, Canada; H16-01648), new patients entering the CCDP from June 2017 until September 2022 were approached for participation in the registry. Data reported in this study represents patients that consented to participate in the registry. During the period of January 1, 2019, to June 21, 2019, recruitment to the Data Registry paused intake of new patients while it redesigned its model of care. From March 12, 2020, until Feb 20, 2021, recruitment was paused due to the COVID-19 pandemic. During this time, major changes to clinical processes were implemented on BC Women’s Hospital campus, including telehealth changes and physical distancing requirements.

### Data Collection

Demographic information was collected before a patient’s first in person appointment at the CCDP. Data points included age, geographical location, sex, gender, ethnicity, marital status, education, employment, and income. Clinical variables are transferred from the CCDP Data Registry from the Interprofessional Assessment (IP) Tool which is completed collaboratively by clinical staff including physicians, occupational therapists, nurses, and social workers. The IP Tool contains data about CCD diagnostic instruments [2,14,15,20], clinical variables, including health history, symptom presentation, functional status, and physical examinations.

The CCDP standardized questionnaires include a range of patient health outcomes:

1. The Fatigue Severity Scale is a nine-item questionnaire that measures the impact of fatigue on the patient’s daily living [21].
2. The Short Form McGill Pain Questionnaire captures the sensory and affective dimensions of subjective pain felt by the patient [22].
3. The Pittsburgh Sleep Quality Index (PSQI) assesses nighttime sleep problems and sleep quality [23].
4. The RAND 36 Short Form Health Survey (SF-36) is an indicator of overall health status based on eight scaled scores. Two component scores, a mental health and physical health component score can be generated to summarize overall physical and mental wellbeing. These scores are reverse coded, wherein a larger score indicates a higher level of wellbeing [24].
5. Personalized Health Questionnaire 9 (PHQ-9) and Generalized Anxiety Disorder 7 (GAD-7) are used to measure patients’ emotional well-being, namely, the presence and severity of depression and anxiety [25,26].
6. The Phenotyping Questionnaire Short Form (PQ-12) is derived from the UK ME/CFS Biobank Participant Phenotyping Questionnaire and is used to assess presence and severity of multiple symptom domains for patients with ME/CFS [27]. Higher scores are associated with higher severity.

The standardized questionnaires were gathered from participants at four time points: Upon entry to the Program, 6 months into the Program, upon discharge (typically after 1 year), and 3-6 months post-discharge. All data was stored in the Research Electronic Data Capture (REDCap) platform hosted by the BC Children Hospital Research Institute (BCCHRI). [28]. Inventory sub-scales and global scores for standardized questionnaires were calculated (using REDCap calculated fields based on the instrument’s cited calculation methods). Scores were not included if key data used to calculate summary scores of the instruments were missing.

### Analysis Strategy

To maintain analytical robustness and ensure sufficient statistical power, some demographic variables were sub-grouped, resulting in lower number of sub-groups. To characterize the patient population, descriptive statistics of collected variables were performed. For continuous variables, the means and standard deviations were calculated, or medians and interquartile ranges (IQR) as appropriate. For categorical variables, counts and proportions were calculated. For longitudinal presentation of each symptom-related variable the sample mean was plotted at each timepoint to display overall trends.

To evaluate the differences in various self-reported symptom variables across different time points – specifically, between baseline and 6-months, and between baseline and discharge – we employed paired samples t-tests. The number of individuals contributing information at each timepoint was displayed in generated tables, allowing for extrapolation of the loss-to-follow up in our study. Normality assumptions were checked using diagnostic Q-Q plots and found satisfactory for our sample [29].

#### Disease Duration

To determine whether disease duration at recruitment influences patient outcomes, two sets of univariable and multivariable linear regressions were modeled. The first tested for associations between disease duration and symptom variable scores at baseline. The second looked at potential associations between disease duration and the difference in symptom scores between baseline and discharge. The decision to include confounders in any of the adjusted models was based on statistical or theoretical significance, with lower Akaike Information Criterion (AIC) values serving as the benchmark for statistical significance [30]. For all linear regression models included, assumptions of linearity, normality, homoscedasticity, and independence were checked and were void of major violations.

#### Model of Care

The CCDP clinical program switched program delivery models from 1:1 consultation with doctors and other clinicians to a predominantly group-based model with allied health professionals in 2019. To characterize differences in health outcomes for the two CCDP models, dummy variables were created to dichotomize participants into two assessment groups. Participants who had an assessment date on or before January 17, 2019, we considered part of the “individual-based” program and those assessed after January 17, 2019, were considered part of the “group-based” program. Two Sample T-Tests were used to compare the difference in symptom scores from baseline to 6-months follow-up across the two groups. The number of individuals with information at “discharge” were too low for this analysis in the individual-based group (n=35), so we restricted the analysis to baseline and 6-months only.

A two-sided p-value <0.05 was considered statistically significant and outcome means were presented with the corresponding 95% confidence intervals. Data were analyzed using R 3.2.3 software (R Foundation for Statistical Computing, Vienna, Austria).

## Results

### Recruitment Metrics

CCDP Data Registry participation was offered to 1962 CCDP patients, 1409 of whom expressed interest in learning more about the study. Of these, 735 patients consented to participate in the study. Finally, 668 participants were included in this analysis. Reasons for exclusion included not meeting diagnostic criteria for the patient population in the CCDP clinic, not completing their Standardized Questionnaires prior to their first appointment (a study exclusion criteria), and withdrawals. Overall, 34% of patients approached for participation were involved in the study. Figure 2 shows a flow chart of study recruitment metrics. Further reductions in sample sizes occur for some analyses due to non-completion of certain questionnaires or loss-to-follow-up.

**Figure 2.**
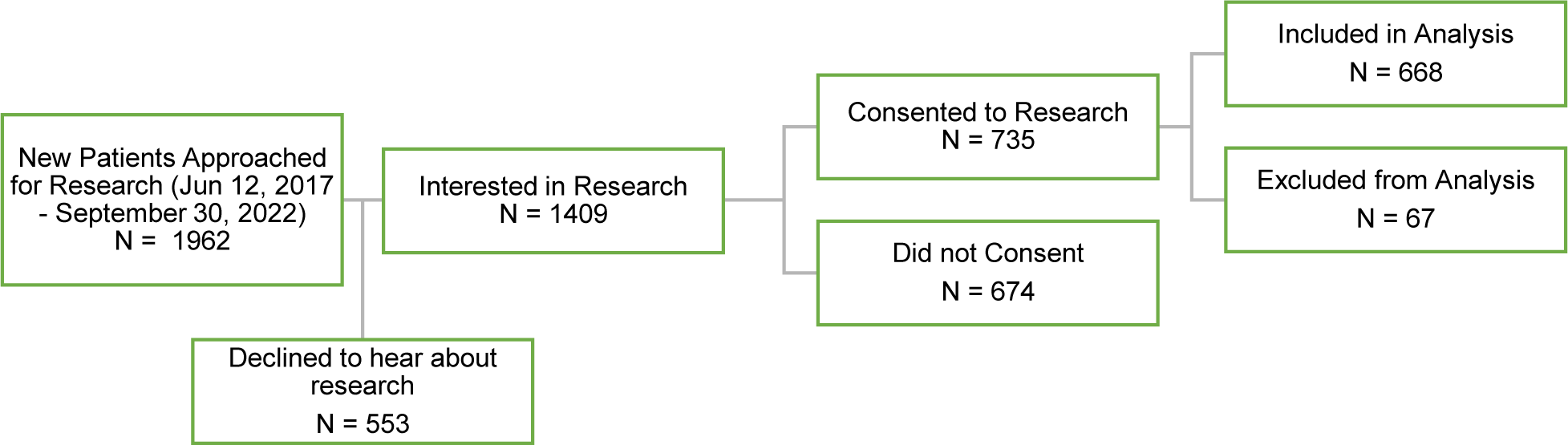
Flowchart of the participants included and excluded at different stages of the CCDP Data Registry recruitment.

### Baseline Characteristics

Table 1 shows a breakdown of the demographic features of the sample at baseline. Diagnoses of ME/CFS and FM were registered by the attending physician. Patients with a diagnosis of ME/CFS encompassed 85% of the sample (n = 568). The number of participants with a diagnosis of FM summed to 460 (69%), with 360 of these participants receiving both diagnoses of ME/CFS and FM. Only two participants had a confirmed diagnosis of CLLS, one of them also had a diagnosis for ME/CFS and FM and was categorized as such, the other was removed from analyses.

**Table 1:**
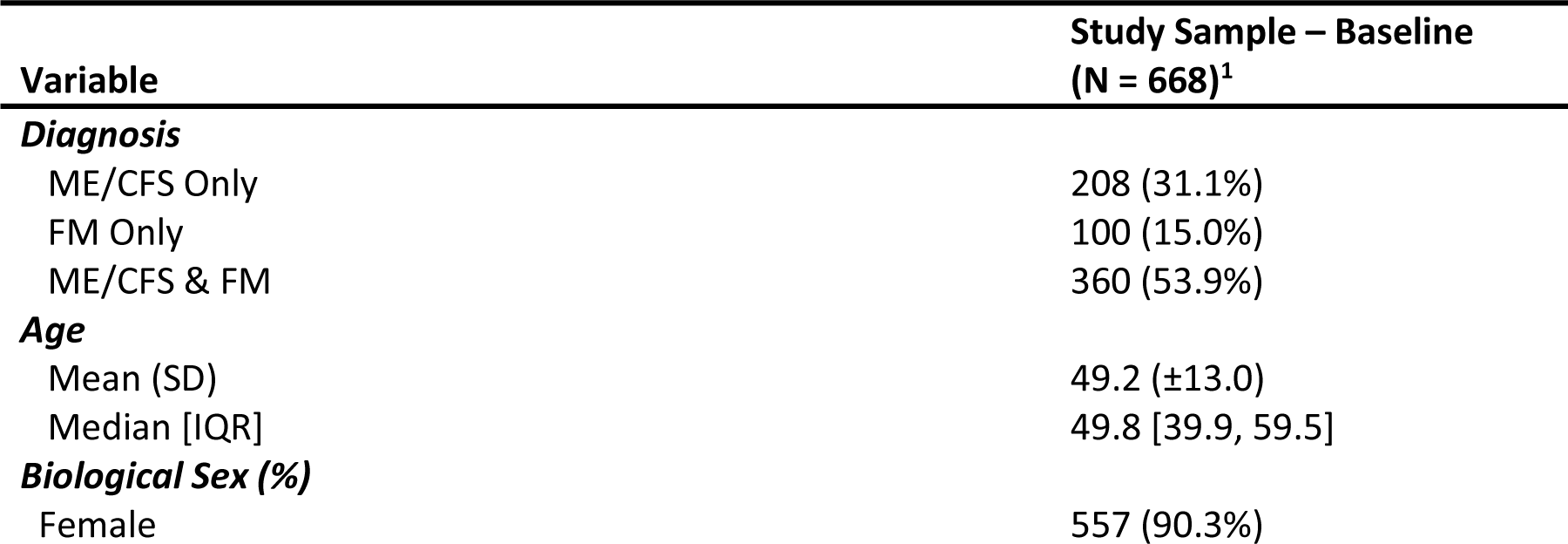
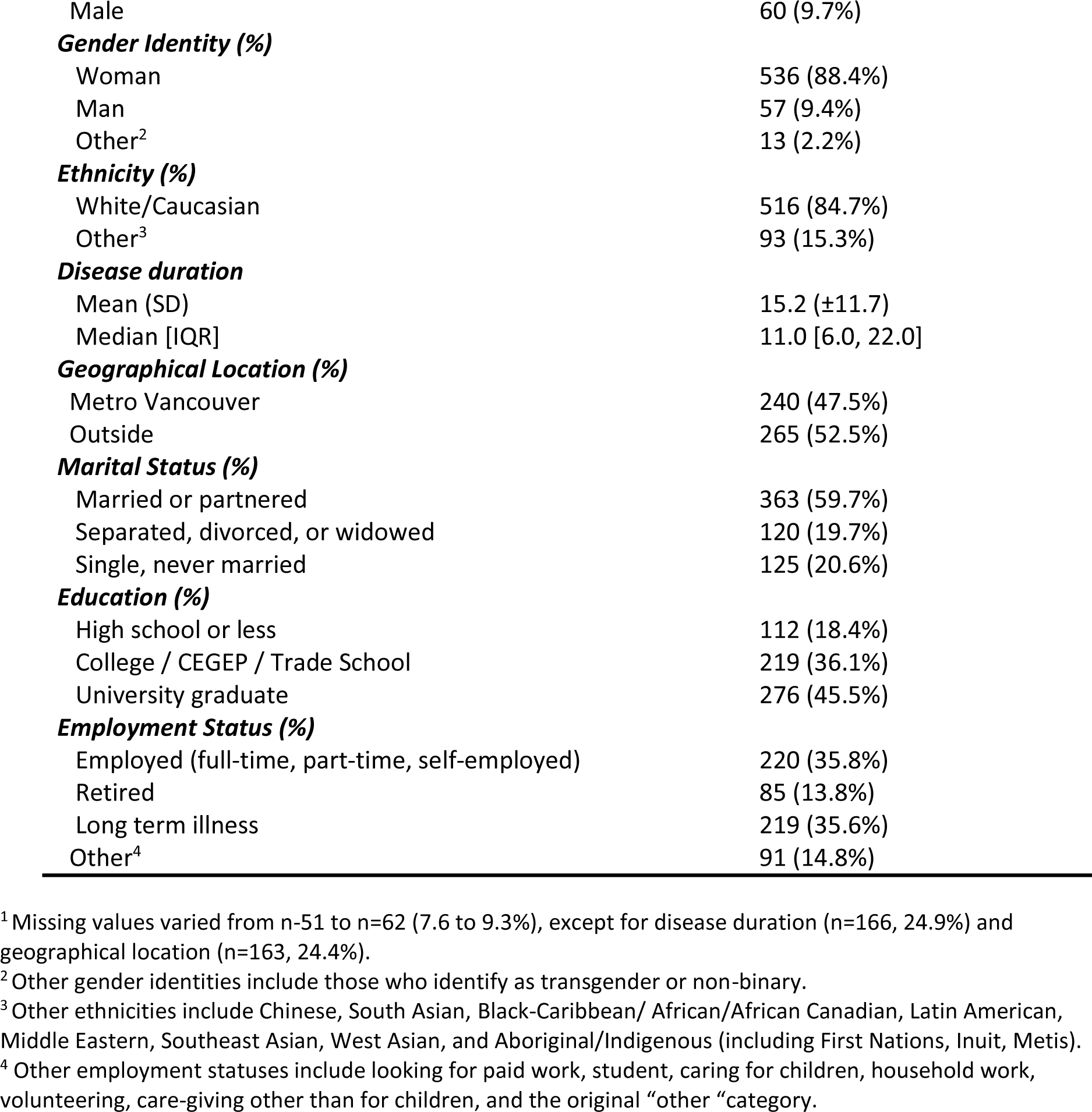
Baseline participant characteristics: CCDP adult patients enrolled in the Data Registry.

| Variable | Study Sample – Baseline<br>(N = 668)<sup>1</sup> |
| --- | --- |
| **Diagnosis** |  |
| ME/CFS Only | 208 (31.1%) |
| FM Only | 100 (15.0%) |
| ME/CFS & FM | 360 (53.9%) |
| **Age** |  |
| Mean (SD) | 49.2 ($\pm 13.0$) |
| Median [IQR] | 49.8 [39.9, 59.5] |
| **Biological Sex (%)** |  |
| Female | 557 (90.3%) |
| Male | 60 (9.7%) |
| **Gender Identity (%)** |  |
| Woman | 536 (88.4%) |
| Man | 57 (9.4%) |
| Other<sup>2</sup> | 13 (2.2%) |
| **Ethnicity (%)** |  |
| White/Caucasian | 516 (84.7%) |
| Other<sup>3</sup> | 93 (15.3%) |
| **Disease duration** |  |
| Mean (SD) | 15.2 (±11.7) |
| Median [IQR] | 11.0 [6.0, 22.0] |
| **Geographical Location (%)** |  |
| Metro Vancouver | 240 (47.5%) |
| Outside | 265 (52.5%) |
| **Marital Status (%)** |  |
| Married or partnered | 363 (59.7%) |
| Separated, divorced, or widowed | 120 (19.7%) |
| Single, never married | 125 (20.6%) |
| **Education (%)** |  |
| High school or less | 112 (18.4%) |
| College / CEGEP / Trade School | 219 (36.1%) |
| University graduate | 276 (45.5%) |
| **Employment Status (%)** |  |
| Employed (full-time, part-time, self-employed) | 220 (35.8%) |
| Retired | 85 (13.8%) |
| Long term illness | 219 (35.6%) |
| Other<sup>4</sup> | 91 (14.8%) |

<sup>1</sup> Missing values varied from n=51 to n=62 (7.6 to 9.3%), except for disease duration (n=166, 24.9%) and geographical location (n=163, 24.4%).
<sup>2</sup> Other gender identities include those who identify as transgender or non-binary.
<sup>3</sup> Other ethnicities include Chinese, South Asian, Black-Caribbean/ African/African Canadian, Latin American, Middle Eastern, Southeast Asian, West Asian, and Aboriginal/Indigenous (including First Nations, Inuit, Metis).
<sup>4</sup> Other employment statuses include looking for paid work, student, caring for children, household work, volunteering, care-giving other than for children, and the original "other" category.

The average age of participants was 49 years old (SD = 13). A significant majority, 90% (n=557) of the sample was female, and 85% (n=516) self-identified as White/Caucasian. Just over half of the participants (n=265) resided outside Metro Vancouver (53%), 60% were married or partnered, and 82% had attended some form of post-secondary education. Employment status showed that 64% of the participants were not engaged in formal employment.

Figure 3 shows the mean health outcome scores at three time points, including all individuals who provided information at that point. They show an overall trend for improvement in outcomes, particularly from baseline to discharge. This figure displays broader trends in the outcome scoring, with the scale differing in each pane. For SF-36 scores, higher values indicate better health status according to the indicator, the opposite is true for all other indicators in the Figure (i.e. lower values meaning lower severity or better health).

**Figure 3.**
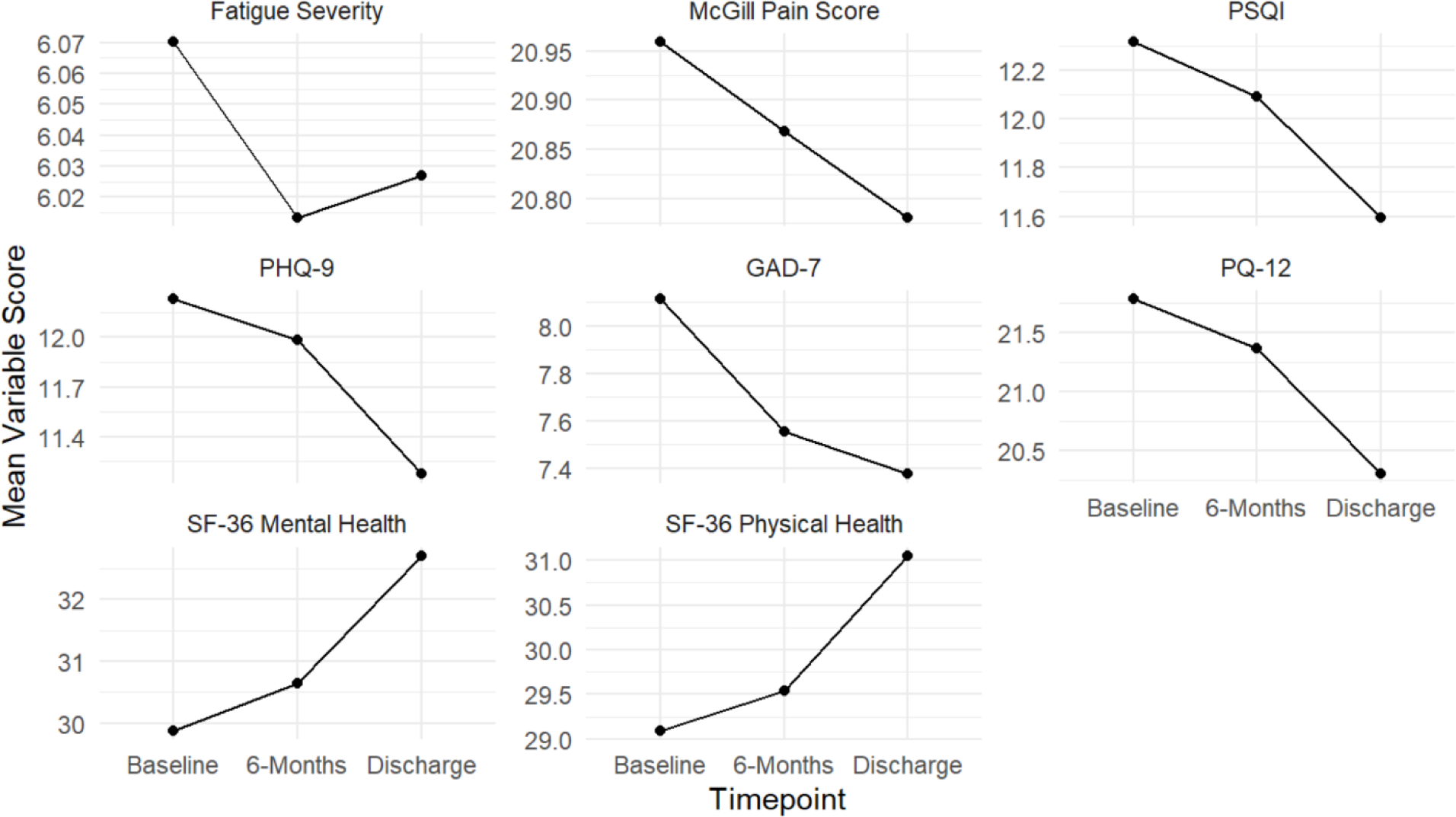
Average symptom variable scores across CCDP follow-up timepoints.

Table 2 displays the mean difference in scores for each health outcome and the results of paired sample t-tests comparing baseline measurements to those at 6 months and discharge. Measures of pain, fatigue and sleep showed overall trends for improvement, but no significant differences between baseline and either follow-up time point. However, overall physical health, represented by the SF-36 Physical Health Summary Score indicated significant improvement by discharge. The PQ-12 showed a significant reduction from baseline to discharge, indicating a reduction in ME/CFS-specific symptoms over time. Regarding emotional well-being, the GAD-7, but not the PHQ-9 presented significant reductions from baseline to both 6 months and discharge, while the SF-36 Mental Health Summary scores showed a significant increase at discharge (p-value < 0.05). Both changes indicate an improvement in mental health over the course of the Program.

**Table 2.**
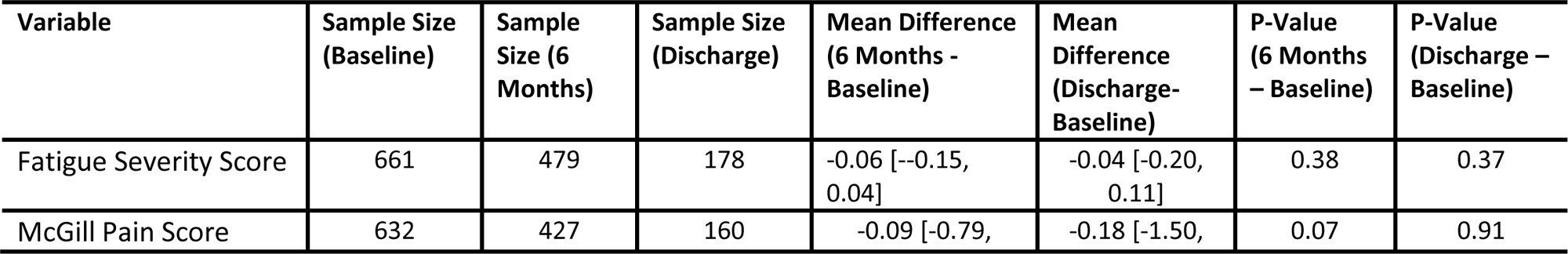
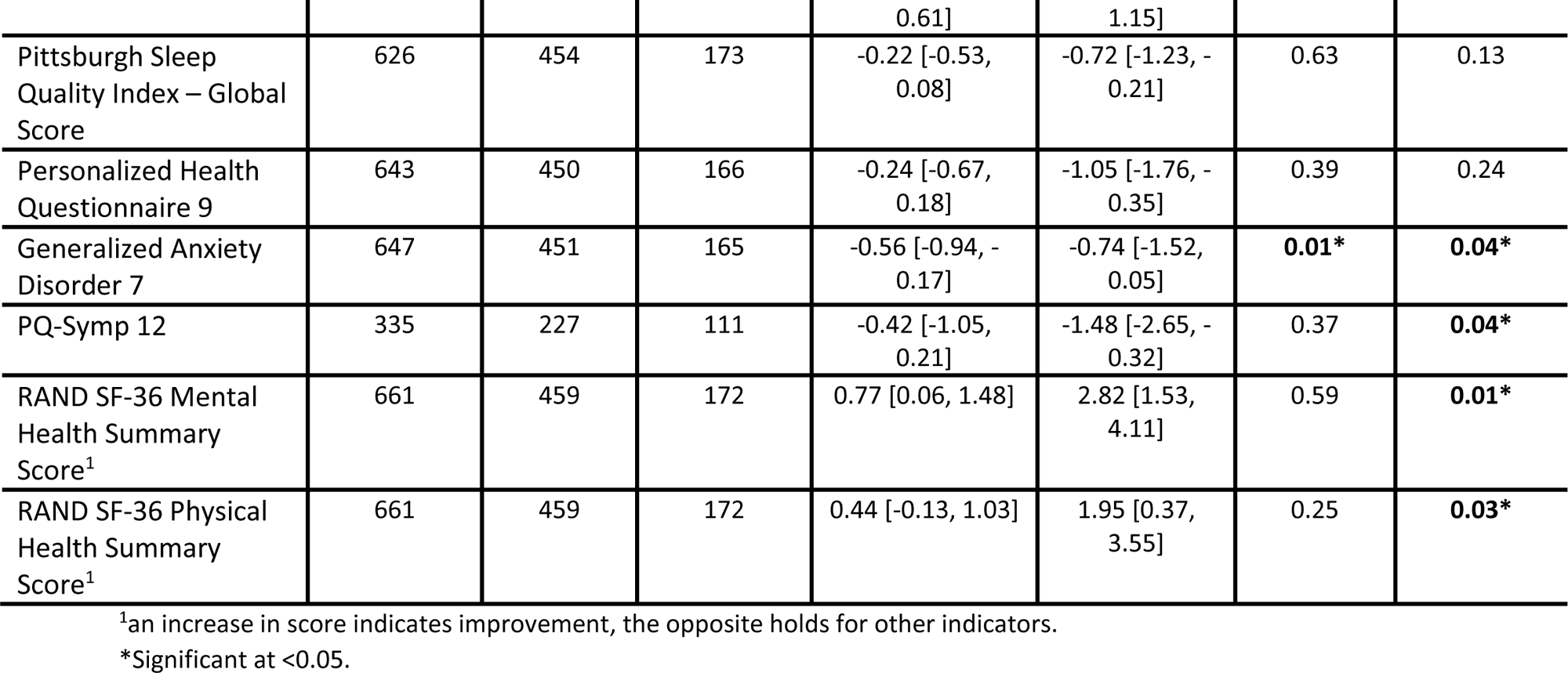
Mean symptom score differences between baseline to 6-month follow-up and baseline to discharge (paired-analysis).

#### Disease Duration

The relationship between disease duration and symptom variables at baseline summarized in Table 3, showed a general trend for more severe scores at baseline in those with higher disease duration at the time they are recruited into the Program. McGill Pain Score and PSQI score are the only significant associations with disease duration in univariable modelling (p<0.05), both indicating worse scores associated with longer disease duration at baseline. These measures were attenuated after adjusting for demographic factors in the multivariable models. The PSQI was the only health-related variable to exhibit a significant association with disease duration (β = 0.033, p = 0.038), where the association persisted in the multivariable model (β = 0.035, p = 0.041). For each additional year of disease, patients would experience a 0.035-point increase in their baseline sleep scores (PSQI), which corresponds to worsening overall sleep quality.

**Table 3.**
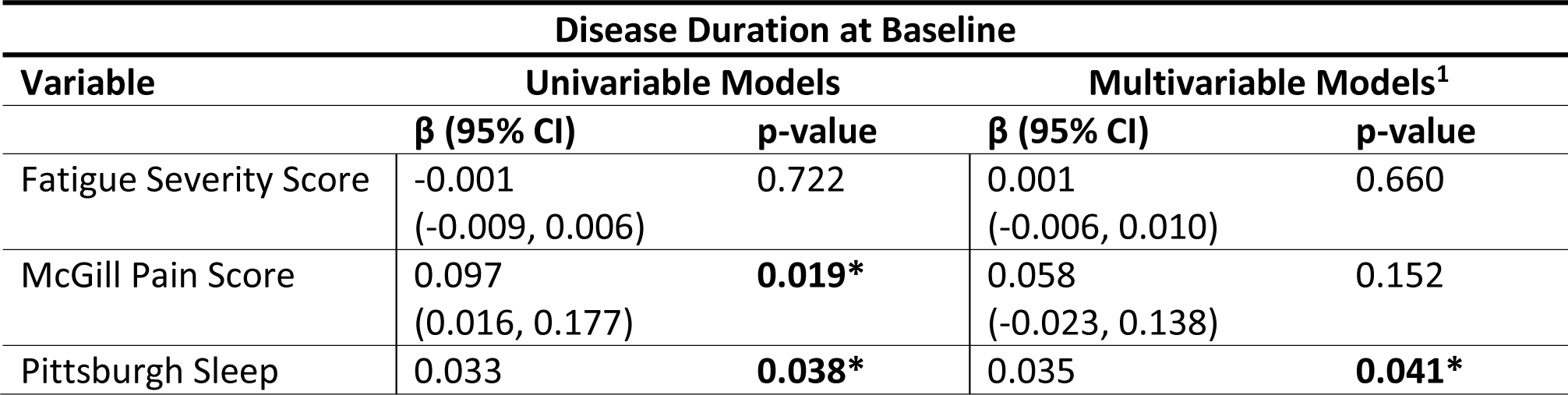
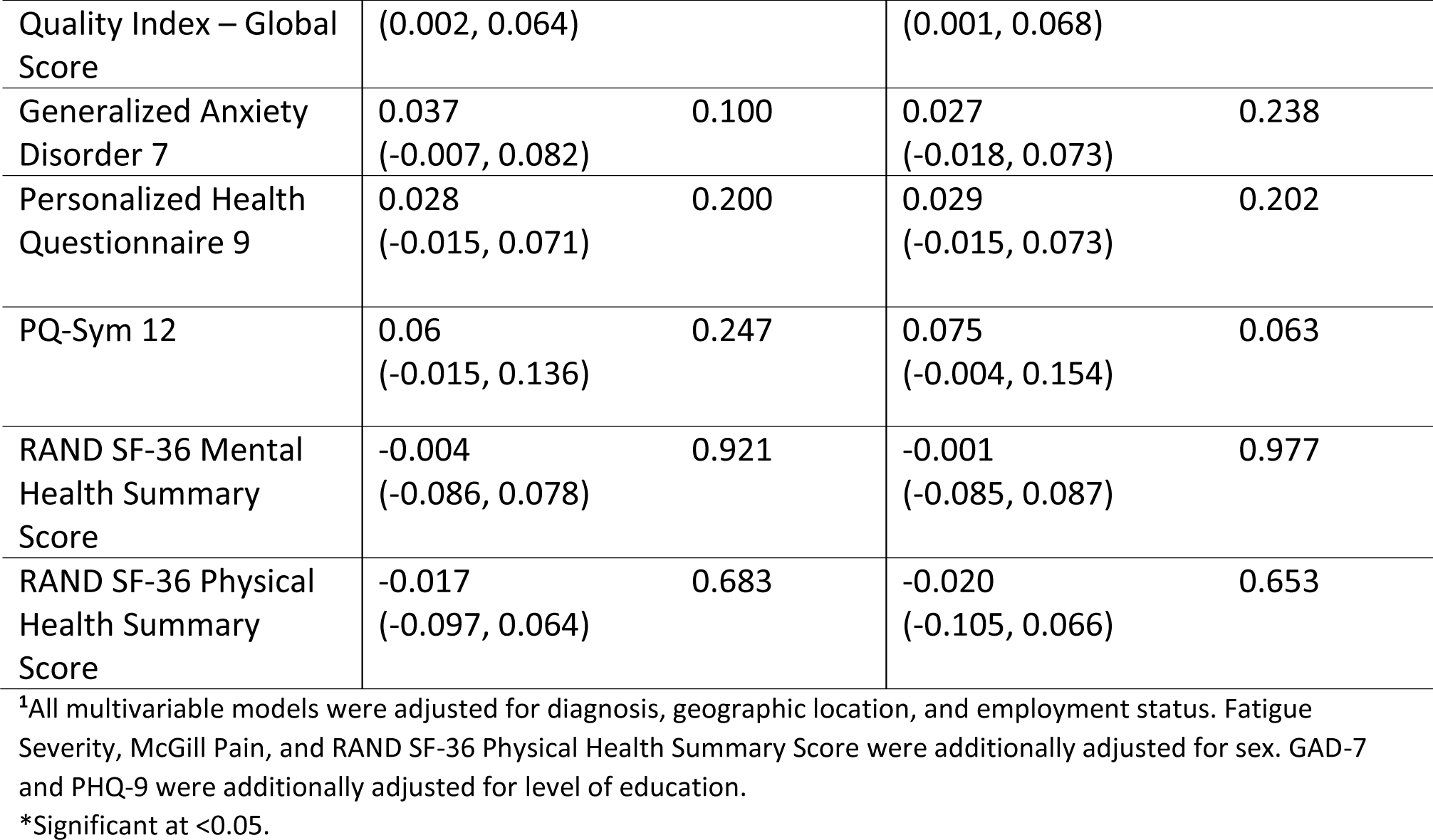
Linear regression models for symptom severity by baseline disease duration (n= 480).

The relationship between disease duration at baseline and symptom variable differences (discharge – baseline) were also examined, but no symptom variable changes retained a significant association after adjustment (supplementary table 1).

#### Model of Care

Patients were categorized as either taking part in the “individual-based” or “group-based” model of care based on their assessment date. When the difference between baseline and 6-month scores were compared across the two groups, we saw that some of the mental health metrics are significantly different (Table 4). Those who took part in the individual-based model of care experienced a greater reduction in both anxiety (GAD-7) and depression (PHQ-9) severity, or around a 1.4-point decrease in scores from baseline to 6-months follow-up. For other scores, the “individual-based” model showed more favourable trends towards improvement, which were not significant.

**Table 4.** Mean difference in scores for standardized health outcome variables across two program models (consultation and group-based). P-values presented for t-tests comparing baseline to 6-months follow-up.

|  | Individual-Based<br>(6-Months – Baseline)<br>(n=155) | Group-Based<br>(6-Months– Baseline)<br>(n=276) | P-Value |
| --- | --- | --- | --- |
| Variable | Mean Diff (SD) | Mean Diff (SD) |  |
| Fatigue Severity Score | -0.13 (0.93) | 0.01 (1.06) | 0.131 |
| McGill Pain Score | -1.56 (8.00) | -0.20 (6.74) | 0.087 |
| Pittsburgh Sleep<br>Quality Index – Global<br>Score | -0.15 (3.31) | -0.04 (3.17) | 0.731 |
| Generalized Anxiety Disorder 7 | -1.46 (3.98) | 0.00 (4.06) | **<0.01\*** |
| Personalized Health Questionnaire 9 | -1.44 (4.08) | 0.52 (4.59) | **<0.01\*** |
| RAND SF-36 Mental Health Summary Score | 0.57 (5.30) | -0.02 (8.73) | 0.371 |
| RAND SF-36 Physical Health Summary Score | 1.01 (6.06) | -0.02 (6.36) | 0.090 |

\*Significant at <0.05.

## Discussion

This analysis of the CCDP Data Registry data offers novel insights into symptom progression of complex chronic diseases under a clinical program, and vast opportunities for continued exploration. Notably, the study documented general trends towards symptom improvement over the course of CCDP enrollment and significant improvements in some mental health, physical health, and ME/CFS -specific symptoms. Individual-based care models demonstrated greater effectiveness in reducing anxiety and depression symptoms compared to group-based models. However, our results revealed a concerning severity and persistence of symptoms among participants over time. The study highlighted the long-term nature of these diseases, with no significant relationship found between disease duration at entry to the Program and symptom severity, except for sleep quality which tended to deteriorate with longer disease duration. This points to the complex challenge of CCD management and future research needs, further emphasized by the modest changes observed despite the CCDP’s comprehensive care approach.

The demographic breakdown of the CCDP Data Registry population showed a composition somewhat consistent with other studies of CCDs. Nearly 90% of the sample reported their biological sex as female and 88% self-identified as women as their gender. This is consistent with other research that reported high proportions of ME/CFS patients being women, between 72-82% [31–34]. In a meta-analysis from 2020, the authors reported prevalence estimates for ME/CFS of 1.36% for females and 0.86% for males [35]. The majority of studies also report between 80-90% of FM cases occurring in women [36–38].

CCDs affect people of all races and ages. In the CCDP Data Registry, however, most patients report being Caucasian/white (85%), compared to 65.6% in the census population for British Columbia [39]. Likewise, the average age in the CCDP Data Registry is 49 years, older than the census population mean for British Columbia of 43 [39]. In the literature, most people are diagnosed with FM during middle age (40 to 60 years) [40], while there are two peaks in incidence for ME/CFS, one in adolescence (10 to 19 years) and another in adulthood (30 to 39 years) [41]. CCDP intake does not represent incident cases as patients have typically had their symptoms or diagnoses for many years prior to entering the Program. Furthermore, it does not include children in the sample as the CCDP only serves adults over 18 years. We grouped the 15% of individuals who did not self-identify as white in our sample to perform analyses. However, in doing so, this process lost granularity. Despite the over-representation of white, middle-class individuals in clinical populations of ME/CFS, population-based research suggests that the illness may be more common among people of lower socioeconomic and minority cultural or ethnic groups [13,42–44]. These discrepancies suggest potential differential health access and/or willingness to participate in research, warranting further investigation and efforts.

The results highlight the severity and persistence of illness and impairment faced by those with CCDs. Self-reported metrics for pain, fatigue, and mental health measures are all substantially worse compared to the healthy population. For example, the Fatigue Severity Scale yields a score between 1 and 7, with a cut-off score of 4 often used to indicate clinically significant fatigue [45,46]. CCDP Data Registry participants have an average FSS score of 6, indicating most participants are severely fatigued across timepoints. Given the discrepancy between the well-being of individuals with CCDs and the general population, and even those with other chronic conditions [47,48], there is a clear need for interventions that are more finely tuned to the nuances of these conditions.

Our findings suggest a general trend in symptom improvement, including statistically significant changes in overall physical health, mental health related indicators, and severity of ME/CFS-specific symptoms, between baseline and discharge. These changes, while modest, highlight that engagement within the CCDP results in slight improvements to overall physical and emotional state. Such findings could be linked to the educational and supportive aspects of the Program, as well as the use of pharmaceutical approaches for symptoms management, which are tailored according to individual needs and preferences. The interaction with health professionals, and the opportunities and resources given to patients for the management and navigation of symptoms, workforce involvement, and healthcare systems may offer an opportunity to empower patients, providing a sense of autonomy over their disease management as well as a sense of community.

Findings did not reveal a relationship between disease duration at entry into the Program and baseline symptom scores, nor their trajectory between baseline and discharge. The only exception was PSQI scoring which indicates that longer disease duration is related to reduced sleep quality at baseline. Though symptom improvement is probable over time and with treatment, there are no single curative treatments for CCDs, and the prognosis for recovery from ME/CFS and FM is low [18,49–51], though more favourable in the initial stages of disease, especially, in the first year [52]. Studies have been inconsistent, but generally an increased disease duration for CCDs has been associated with poorer health outcomes [53–56]. We may not see the impacts of greater disease duration in our study because most of our participants have had their CCDs for many years prior to joining the Program (mean disease duration = 11 years), so major declines that may occur in the early stages of disease were not observed. It is also possible that participation in the Program selects against those with more favourable outcomes at earlier disease stages, as well as those very severely affected, contributing to the limitations in interpreting findings on the impact of timing of treatment on disease outcomes.

Our analysis indicated that the difference in health outcomes from baseline to 6-months for the two models of care only differed significantly for mental health outcomes, in spite of some apparent overall advantages of the 1:1 model of care. The transition from a 1:1 individual-based consultation model of care to a group-based program was undertaken to reduce the waitlist time for the CCDP and ensure more patients could receive care. On average, patients in the 1:1 group experienced a greater reduction in anxiety and depression at 6-months compared to the group-based patients. This may be due to the more personalized support provided. However, it is important to consider factors external to the CCDP, a significant example being that most of those doing group activities, did so during the COVID pandemic, where pressures on mental health, and overall access to services were noted across society. As such, it is possible that the ongoing burden of a concurrent pandemic may have resulted in some inflation of mental health scoring. Future research could benefit from controlling for external and confounding factors, either through design or analysis.

The CCDP stays abreast of literature and guidance on treatment and care strategies for people with CCDs. However, there is limited evidence for effective interventions that are robust and reproducible [57,58]. Studies on group-based self-management programs for ME/CFS and FM show mixed outcomes and levels of effectiveness [59–61]. Our results show that the CCDP experienced similarly mixed and modest improvements for CCD health outcomes. New approaches and research are urgently needed for therapeutic interventions for CCDs.

Individuals who remain ill after SARS-CoV-2 infection are labeled with Post-Acute Sequelae of SARS-CoV-2 Infection (PASC) and share similar symptom profiles and features to patients with ME/CFS [62,63]. The parallels between PASC and ME/CFS, particularly post-infectious ME/CFS, may suggest a shared pathophysiology. This presents an opportunity for synergistic research efforts into understanding post-viral syndromes and identification of diagnostic markers and therapeutic targets for CCDs.

### Strengths and Limitations

The strengths of this study include the large sample size at baseline and numerous data types collected, encompassing a range of variables that go beyond those analyzed in this paper. The repeated measures design allows for tracking patient journeys individually and in aggregate throughout the Program. The cohort was also well defined, the diagnosis of ME/CFS and Fibromyalgia were confirmed clinically and entered into the database by a trained practitioner, minimizing misclassification and missingness. Since the CCDP is a provincial referral centre, findings can be cautiously applied to the larger context of BC adults with CCDs, though the sample is less diverse than the BC population.

The sample lacks the diversity of the general population and is limited by its relatively small sample size at discharge due to high attrition and the ongoing data collection for participants currently enrolled in the Data Registry. Iindividuals who dropped out of the study were not completely random, and although efforts were made to adjust for this differential attrition bias in our analysis, we cannot rule out the possibility of it biasing the findings. Measures of health-related variables in this study were assessed using self-reported questionnaires, subject to recall bias and often less sensitive than objective measures. Additionally, the lack of a control group with individuals in the community, but not in the Program, limits the interpretability of our findings. As noted above, the COVID-19 pandemic and other external factors may have had an independent impact on the health of the population, including on the ability to recover from chronic diseases, such as those cared for at the CCDP.

A significant challenge for researching CCDs is appropriate case assessment due to the absence of diagnostic biomarkers. While CCDP patients are identified using established diagnostic protocols, the symptom presentation of CCDs are non-specific, making it difficult to assure a standard phenotype or to distinguish these diseases from others with similar presentations [64,65]. This reliance on clinical diagnoses, in the absence of biomarkers, introduces an element of variability that future research must address.

### Future of the Data Registry

The future state of standardized data collection in clinical centres servicing patients with CCDs is in progress. In addition to the CCDP Data Registry transforming into the ICanCME Network Clinical Cohort, it will include standardization of data collection, harmonized data sets, and a data access portal to enable further research. Additional developments, subject to further funding, include optimization of the follow-up processes to increase response rates and the inclusion of a waitlist control group of individuals waiting to be seen by their regional clinic.

As the only centre for CCDs in Western Canada, the CCDP is intended to be iterative and innovative, piloting and introducing new forms of therapeutics and self-management strategies. The CCDP Data Registry provides a tool to determine whether these approaches are making meaningful improvements in patient’s symptoms and overall health, and to act as a repository for researching this understudied population. This data serves as a valuable benchmark for future data analysis as the CCDP continues to evolve.

## Conclusion

Our findings suggest that patient participation in the CCDP is associated with some improvement in symptoms by discharge for physical and mental health measures. We did not find consistent evidence for improvement in specific symptoms such as pain, sleep quality, and fatigue over the course of treatment at the CCDP. Longer disease duration corresponded to poorer sleep quality at baseline but no changes in symptom progression throughout the Program. The previous individual-based model of care showed greater improvements for mental health than the current group-based model of care, however, for most outcomes, the results were comparable between models of care. The Data Registry shows its potential for evaluating interventions and health outcomes in clinical populations with CCDs.

## Data Availability

De-identified data produced in the present study may be available upon reasonable request to the authors

## Acknowledgments

The Principal Investigator (PI) wishes to extend gratitude to the Pacific Public Health Foundation (previously known as the BCCDC Foundation for Public Health) for their generous funding support of the CCDP Data Registry from April 1, 2018, to March 31, 2023. and for their sponsorship of the PI’s protected research time through the Research Scholar Award from April 1, 2023, to March 31, 2024. In addition, Dr Nacul is grateful for the support from the BCW Health Foundation for supporting his research. We thank the CCDP and WHRI staff who contributed with their expertise to the various stages of development of the Program, facilitation of data collection and delivery of interventions for and with the patient community. We extend gratitude to Sabina Dobrer, the Senior Statistician at the WHRI for their expert advice on statistical methods and review of the manuscript’s data analysis section. We are grateful to all those within and outside the Program, including patients and knowledge users who contributed to the Data Registry and the Strategic Direction Plan.

