## Supplemental Table 1 for "Complex Chronic Diseases Program: Program Description & Health Outcomes Assessment from a Clinical Data Registry"

### Supplementary Information

**Supplementary Table 1.** Linear regression models for differences in health outcome scores (discharge – baseline) with disease duration at recruitment as the explanatory variable.

| Disease Duration at Baseline | | | | |
|---|---|---|---|---|
| **Variable** | **Univariable Models** | | **Multivariable Models[1]** | |
| | **β (95% CI)** | **p-value** | **β (95% CI)** | **p-value** |
| Fatigue Severity Score | 0.006 (-0.007, 0.020) | 0.384 | -0.001 (-0.016, 0.015) | 0.946 |
| McGill Pain Score | 0.121 (0.003, 0.239) | **0.044*** | 0.097 (-0.039, 0.234) | 0.159 |
| Pittsburgh Sleep Quality Index – Global Score | 0.010 (-0.039, 0.058) | 0.694 | 0.001 (-0.053, 0.056) | 0.964 |
| RAND SF-36 Physical Health Summary Score | -0.066 (-0.221, 0.080) | 0.405 | -0.027 (-0.214, 0.161) | 0.779 |
| RAND SF-36 Mental Health Summary Score | -0.107 (-0.228, 0.015) | 0.085 | -0.119 (-0.263, 0.025) | 0.104 |
| Generalized Anxiety Disorder 7 | 0.002 (-0.077, 0.082) | 0.952 | 0.026 (-0.058, 0.111) | 0.536 |
| Personalized Health Questionnaire 9 | -0.006 (-0.074, 0.062) | 0.864 | -0.016 (-0.095, 0.064) | 0.698 |
| PQ-12 | -0.031 (-0.143, 0.081) | 0.586 | -0.050 (-0.179, 0.079) | 0.441 |

[1]All multivariable models were adjusted for diagnosis, geographic location, and employment status. Fatigue Severity, McGill Pain, and RAND SF-36 Physical Health Summary Score were additionally adjusted for sex. GAD-7 and PHQ-9 were additionally adjusted for education.

*Significant at <0.05.
